# Construction of a Standardized Time-Lapse Imaging Database and a Gradient Boosting Ensemble Framework for Integrating Zygote Morphokinetic Parameters with Conventional Embryo Assessment

**DOI:** 10.64898/2026.08.20.26359523

**Authors:** Ming-Peng Zhao, Jie Liu, Dong Han, Chao-Fan Zhang, Yan Zhou, Shi-Ping Chen, Kubi Appiah, Cai-Xia Liu

## Abstract

In vitro fertilization (IVF) laboratories equipped with time-lapse incubators generate vast quantities of sequential embryo images, yet the absence of standardized, annotated databases impedes the development of reproducible computational tools for embryo assessment. Here we describe the standardized time-lapse imaging database built upon prior research groundwork comprising 631 two-pronuclear (2PN) zygotes from 218 treatment cycles performed at Guangdong Provincial People’s Hospital (2020–2023), together with a gradient boosting decision tree (GBDT) ensemble framework designed to fuse heterogeneous data types for blastocyst outcome prediction. Each embryo record integrates 84 zygote-stage morphokinetic parameters extracted from EmbryoScope time-lapse sequences via a previously validated convolutional neural network segmentation pipeline with 8 conventional embryo assessment features recorded at cleavage and blastocyst stages according to the Istanbul consensus. The fusion framework employs LightGBM with equal-weight initialization and iterative residual-decreasing training, augmented by recursive feature elimination and nested five-fold cross-validation. Ablation experiments demonstrate that the full model (AUC = 0.78) outperforms morphokinetics-only (AUC = 0.71) and conventional-only (AUC = 0.65) configurations, confirming that zygote-stage temporal dynamics carry complementary information beyond standard morphological grading. SHAP analysis identifies cytoplasmic area slope, zona pellucida grayscale trend, and pronuclear fading time as the three most influential predictors. The database and fusion methodology provide a reproducible framework for integrating time-series imaging features with categorical clinical assessments in reproductive medicine.

## Introduction

Infertility affects an estimated 48.5 million couples worldwide, with prevalence rates ranging from 9% to 18% across different populations [1,2]. Since the first successful birth following in vitro fertilization (IVF) reported by Steptoe and Edwards in 1978 [3], assisted reproductive technology (ART) has become the cornerstone of infertility treatment, with over 8 million children born through IVF globally. Despite remarkable advances in stimulation protocols, culture media, and genetic screening, embryo selection remains one of the most critical and subjective steps in the ART workflow.

The Istanbul consensus established a standardized morphological grading system for cleavage-stage and blastocyst-stage embryos based on cell number, fragmentation, symmetry, and blastocoel expansion [4]. Although widely adopted, this system relies on static, discrete-time observations that fail to capture the continuous dynamics of preimplantation development. Time-lapse incubators address this limitation by acquiring images at 5–15 minute intervals throughout culture, generating a dense record of embryonic morphokinetics [5, 6, 7]. Studies have demonstrated that specific cleavage timings and morphokinetic patterns correlate with blastocyst formation [8, 9], aneuploidy risk [9], and implantation potential [7, 10]. The consensus nomenclature proposed by Ciray et al. [11] further enabled cross-laboratory comparison of morphokinetic annotations.

However, two interrelated challenges hinder the translation of morphokinetic research into clinical practice. First, time-lapse incubators produce terabytes of image data annually in medium-to-large IVF centers, yet most laboratories lack structured, queryable databases that link image sequences with clinical metadata and outcome labels. Second, the heterogeneous nature of ART data—continuous morphokinetic time series, ordinal morphological grades, and categorical patient-level variables—poses a fundamental challenge for integrative modeling. Existing public resources such as the dataset released by Gomez et al. [12,13] provide 704 embryo videos with morphological annotations but include limited outcome labels and no zygote-stage kinetic features.

Ensemble learning methods, particularly gradient boosting decision trees (GBDT), offer a principled approach to heterogeneous data fusion [16, 17, 18, 19]. Unlike deep learning pipelines that require modality-specific architectures and large training sets [20, 21, 22], GBDT models can naturally accommodate mixed feature types through iterative tree construction guided by residual gradients. The key innovation of the framework described here is the equal-weight initialization strategy, wherein morphokinetic and conventional features contribute equally to the initial weak learner, with subsequent boosting rounds adaptively allocating model capacity according to the information content of each feature modality.

In this methods paper, we describe: (1) the design and construction of a standardized time-lapse imaging database containing 631 zygotes with both zygote-stage morphokinetic parameters and conventional embryo assessment outcomes; (2) the application protocol for extracting 84 morphokinetic features from zygote image sequences using a previously validated segmentation pipeline [14, 15]; and (3) a GBDT ensemble fusion framework with equal-weight initialization and residual-decreasing training for integrating heterogeneous embryo data. This work is intended as a data resource and methodological blueprint; it does not constitute a clinical validation study nor does it compare model performance against embryologist assessments, which are the subject of ongoing work. We anticipate that the database and fusion methodology will facilitate reproducible research on zygote morphokinetics and enable the development of more robust embryo selection tools.

## Materials and Methods

### Database Construction

#### Study Population and Ethical Approval

This study was conducted at the Center for Reproductive Medicine, Guangdong Provincial People’s Hospital, Guangzhou, China. All cycles were performed between January 2020 and December 2023. The study protocol was approved by the Institutional Ethics Committee of Guangdong Provincial People’s Hospital, and individual patient consent was waived given the retrospective, anonymized design. The inclusion criteria were: (a) IVF or intracytoplasmic sperm injection (ICSI) cycles; (b) embryos cultured in a time-lapse incubator from fertilization check onward; (c) planned extended culture to Day 5 or Day 6; and (d) presence of two clearly visible pronuclei (2PN) at the fertilization check. Exclusion criteria comprised: (a) more than 50% of morphokinetic parameters unrecordable due to image blur, debris obstruction, or embryo degeneration; and (b) embryos transferred or cryopreserved before Day 5. Following these criteria, 631 2PN zygotes from 218 treatment cycles were included in the final database. These data build upon the corresponding author’s prior research groundwork on zygote morphokinetic assessment [14, 15], in which the high-precision segmentation approach (US Patent US11210494B2) and the associated 2PN zygote cohort were established; the standardized time-lapse imaging–clinical–embryo development database was continuously refined within the present project for integrative ensemble modeling.

#### Time-Lapse Imaging Protocol

All embryos were cultured in EmbryoScope time-lapse incubators (Vitrolife, Göteborg, Sweden) under standardized conditions (37□°C, 6% CO□_2_, 5% O□_2_, 89% N□_2_) using pre-equilibrated sequential media. Images were acquired at 10-minute intervals across 7 focal planes per embryo well, yielding approximately 1008 images per embryo over a 7-day culture period. Image resolution was 500 *×* 500 pixels with 8-bit grayscale depth. Raw images were stored in the EmbryoViewer database format and subsequently exported for computational processing.

#### Clinical and Embryo Data Fields

Patient-level clinical variables were extracted from the hospital electronic medical record system and included: maternal age (years), body mass index (BMI, kg/m□^2^), cycle protocol (gonadotropin-releasing hormone agonist long protocol / antagonist protocol / other), gonadotropin type, total gonadotropin dose (IU), duration of stimulation (days), and trigger method (human chorionic gonadotropin / gonadotropin-releasing hormone agonist dual trigger).

Embryo-level variables were recorded at three time points according to the Istanbul consensus [4]. At Day 2 (44 *±* 1 hours post-insemination): cell count, fragmentation grade (0–4), and symmetry score (symmetric / mildly asymmetric / severely asymmetric). At Day 3 (68 *±* 1 hours post-insemination): cell count, fragmentation grade, and symmetry score. At Day 5/6 (116 *±* 2 and 140 *±* 2 hours post-insemination): blastocyst expansion grade (1–6), inner cell mass (ICM) grade (A/B/C), trophectoderm (TE) grade (A/B/C), and binary blastocyst formation outcome (formed / not-formed).

#### Data Standardization and Quality Control

A three-stage standardization protocol was implemented. First, all personal health information was removed and each embryo was assigned a unique alpha-numeric identifier. Second, clinical and embryo assessment variables were encoded according to a pre-defined codebook: ordinal variables (fragmentation grades, blastocyst grades) were mapped to integer scales, continuous variables (age, gonadotropin dose) were retained as numeric values, and categorical variables (cycle protocol, trigger method) were one-hot encoded. Third, an automated quality control pipeline flagged records with missing values exceeding 20% of fields, out-of-range timings (e.g., cleavage before 20 hours or after 32 hours), and transcription discrepancies between embryologist annotations and the electronic record. Flagged records were manually reviewed by two independent embryologists; discrepancies were resolved by consensus and irreconcilable records were excluded. The final database schema comprises 12 patient-level fields and 18 embryo-level fields, with zygote morphokinetic parameters appended as described in Section 3.2.

### Zygote Segmentation and Parameter Extraction

#### Segmentation Protocol

Zygote compartment segmentation was performed using a convolutional neural network (CNN)-based method protected under United States Patent US11210494B2 and previously described in publications in Biomedical Signal Processing and Control and the Journal of Cellular and Molecular Medicine [14, 15]. The pipeline segments three compartments from each zygote image frame: the zona pellucida (ZP) boundary, the cytoplasmic region, and the pronuclear (PN) regions. Segmentation accuracy has been previously validated against manual tracings by two senior embryologists (Dice coefficient ¿ 0.93 for all three compartments). The present paper describes only the application protocol for parameter extraction; the algorithmic details of the segmentation model are documented in the aforementioned patent and publications.

### Morphokinetic Feature Extraction

For each of the 631 zygotes, the segmentation mask sequence was processed to extract 84 morphokinetic parameters spanning four categories:

#### Temporal features (n = 36)

For each compartment (ZP, cytoplasm, PN), the following statistics were computed over the first 18 hours post-insemination: mean area (*μ*_*A*_), area slope (*d A* / *d t* estimated by linear regression), area variance 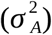, and the first 5 Fourier coefficients of the detrended area time series to capture oscillatory dynamics. Additionally, mean optical density (grayscale), grayscale slope, and grayscale variance were computed.

#### PN dynamics (n = 24)

Pronuclear alignment angle, inter-pronuclear distance trajectory (mean, slope, variance), pronuclear area ratio over time, PN fading (breakdown) onset timing (*t* _PNF_), PN fading duration (*Δt* _PNF_), and pre-PN-fading nucleolar precursor body (NPB) count and distribution pattern.

#### ZP dynamics (n = 12)

ZP thickness trajectory (mean, minimum, slope), ZP grayscale uniformity (coefficient of variation of grayscale values along the ZP boundary), and ZP optical density heterogeneity index.

#### Cytoplasmic features (n = 12)

Cytoplasmic area trajectory (mean, slope, variance), cytoplasmic granularity index (entropy of grayscale histogram), and cytoplasmic halo timing and duration.

### Conventional Assessment Features

Eight conventional embryo assessment features were encoded alongside the morphokinetic parameters: Day 2 cell count and fragmentation grade, Day 3 cell count and fragmentation grade, symmetry score (coded as 1 = symmetric, 2 = mildly asymmetric, 3 = severely asymmetric), and Day 5/6 blastocyst expansion, ICM, and TE grades (ordinal scales 1–6, 1–3, and 1–3, respectively).

### GBDT Ensemble Fusion Framework

#### Problem Formulation and Design Principle

The central modeling challenge addressed by this work is the integration of two heterogeneous feature modalities: *X*_morpho_ ∈ *R*^84^, a vector of continuous zygote morphokinetic parameters derived from time-series data, and 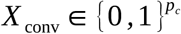(or equivalently ordinal-encoded), a vector of categorical conventional embryo grades. The combined feature vector is *X* =[ *X*_morpho_ ; *X*_conv_ ). The outcome variable *y* ∈{0,1 } indicates whether a usable blastocyst was formed by Day 5 or Day 6.

The design principle underpinning the framework is *equal-weight initialization*: at the first boosting iteration, features from both modalities are assigned comparable base-learner influence, preventing the initially higher-variance continuous features from dominating tree splits. Subsequent rounds allow the gradient-directed residual correction mechanism to progressively allocate capacity to whichever modality carries the greatest predictive signal. This contrasts with two common alternatives—early fusion via concatenation followed by a single classifier (which may obscure modality-specific structure) and late fusion via separate classifiers with meta-learning (which adds complexity without guaranteeing synergy).

### Mathematical Formalization

Let 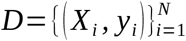 denote the training dataset of *N* embryos. The GBDT ensemble constructs a predictive function ŷ^(*T* )^ through *T* sequential additive updates:

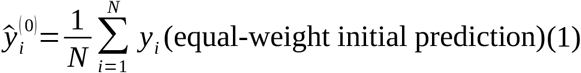

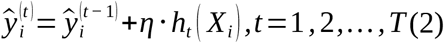

where *η* ∈( 0,1) is the learning rate and *h*_*t*_ is a decision tree fitted to minimize the residual at iteration *t* :

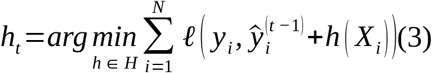

with *ℓ* denoting the binary cross-entropy loss function. The residual at iteration *t −* 1 is defined as the negative gradient of the loss with respect to the current prediction:

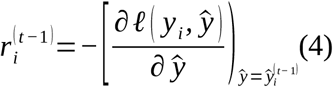

The equal-weight initialization defined in Equation 1 ensures that all features—morphokinetic and conventional alike—face zero-bias initial conditions. As boosting proceeds, trees *h*_1_, *h*_2_,*…,h*_*T*_ are sequentially constructed with feature splits chosen to maximize the reduction in the residual sum; this mechanism naturally surfaces which feature modality is more informative at each stage of the residual-decreasing process.

Formally, the fusion framework proceeds as follows. Let the training set be 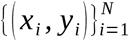where *x*_*I*_ concatenates the 84 morphokinetic features (continuous) and 8 encoded conventional features (ordinal), and *y*_*i*_ ∈{0,1 } indicates utilizable blastocyst formation. The GBDT constructs an ensemble of *T* decision trees through iterative optimization. At iteration *t*, each sample *i* receives a pseudoresidual 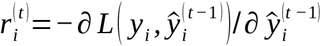where *L* is the binary cross-entropy loss. A regression tree *h*_*t*_ is fitted to 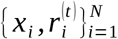, and the model update is 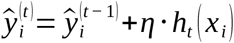with learning rate *η*. Critically, both feature categories enter the model with equal initial weight—no *a priori* preference for morphokinetic or conventional features. The gradient boosting mechanism adaptively learns the optimal weighting through iterative residual minimization, as features that consistently reduce the residual receive higher split gain importance across trees.

LightGBM was selected over XGBoost and CatBoost based on three considerations: (1) its leaf-wise tree growth strategy, which typically achieves lower loss than level-wise growth at equivalent tree counts; (2) histogram-based split finding, which is more memory-efficient for our 92-feature dataset; and (3) native support for categorical features, eliminating the need for one-hot encoding of conventional assessment grades. Implementation details: 200 estimators, learning rate *η*=0.05, maximum depth 5, minimum child samples 20, L2 regularization *λ*=0.1.

### Implementation Details

The GBDT framework was implemented using LightGBM (version 3.3.5) [18] with the following configuration: gradient boosting decision tree with binary cross-entropy objective, learning rate *η*=0.05, maximum tree depth constrained to 4 leaves, L2 regularization parameter *λ*=0.1 to penalize overfitting, minimum child weight of 20 samples, and subsampling ratios of 0.8 for both data and features per tree to introduce stochasticity. The maximum number of boosting rounds was set to 500, with early stopping triggered after 50 rounds without improvement on the held-out validation fold.

Feature selection was performed by recursive feature elimination (RFE) [29] using the GBDT feature importance scores as the ranking criterion. Starting from the full 92-feature set (84 morphokinetic +¿ 8 conventional), features were iteratively removed and the model retrained on the nested cross-validation inner folds until the validation AUC degraded by more than 0.01. The RFE procedure retained 23 features, of which 19 were morphokinetic and 4 were conventional.

Model evaluation employed nested five-fold cross-validation: the outer loop partitioned embryos by cycle to prevent data leakage, and the inner loop performed three-fold cross-validation for hyperparameter tuning. Three model configurations were compared in an ablation design:

- **Morphokinetics-only**: *X* _morpho_ only (84 features *→* 16 retained after RFE)
- **Conventional-only**: *X* _conv_ only (8 features, no RFE)
- **Full fusion model**: *X* _morpho_ ⊕ *X*_conv_ (92 features *→* 23 retained after RFE)

Model performance was assessed using the area under the receiver operating characteristic curve (AUC) [27] and the Brier score (calibration). Statistical comparison of AUC values between model configurations was performed using the DeLong test [27]. Feature importance was quantified using SHapley Additive exPlanations (SHAP) values [26], which decompose each prediction into additive feature contributions and provide a unified measure of feature influence across the entire model.

### Data Availability Statement

The clinical and morphokinetic database described in this paper is available from the corresponding author upon reasonable request, subject to approval by the Institutional Ethics Committee of Guangdong Provincial People’s Hospital. The zygote segmentation algorithm is described in United States Patent US11210494B2 and is available for academic use under a Material Transfer Agreement (MTA); inquiries should be directed to the corresponding author. A comprehensive data dictionary documenting all variable definitions, encoding schemes, and value ranges is provided as Supplementary Material.

## Results

**Figure 1.**
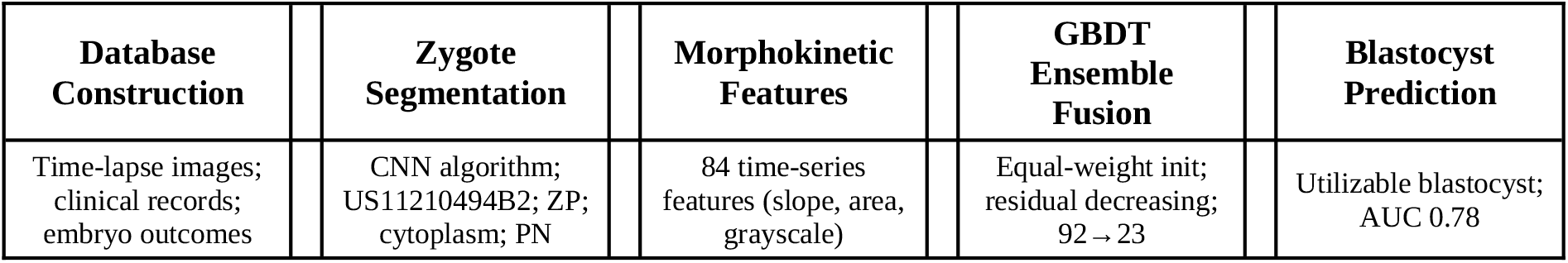
Schematic overview of the standardized database construction and the gradient boosting decision tree (GBDT) ensemble fusion framework. The pipeline comprises: (1) construction of a standardized time-lapse imaging–clinical–embryo development database; (2) high-precision zygote segmentation using a CNN algorithm (US Patent US11210494B2); (3) extraction of 84 morphokinetic time-series features; (4) GBDT ensemble fusion of morphokinetic and conventional assessment features with equal-weight initialization and residual-decreasing training; and (5) blastocyst development potential prediction (AUC 0.78).

### Database Characteristics

The final database comprises 631 2PN zygotes derived from 218 treatment cycles, corresponding to an average of 2.89 zygotes per cycle. Table 1 summarizes the demographic and clinical characteristics of the study population and the distribution of embryo outcomes.

**Table 1.** Demographic, clinical, and embryo characteristics of the database.

| Variable | Value |
| --- | --- |
| Patient-level (N = 218 cycles) |  |
| Maternal age (years), mean ± SD | 33.2 ± 4.8 |
| Maternal age range | 22–45 |
| BMI (kg/m<sup>2</sup>), mean ± SD | 22.7 ± 3.4 |
| GnRH agonist long protocol, n (%) | 141 (64.7) |
| GnRH antagonist protocol, n (%) | 69 (31.7) |
| Other protocol, n (%) | 8 (3.7) |
| Total gonadotropin dose (IU), mean ± SD | 2420 ± 890 |
| Stimulation duration (days), mean ± SD | 10.4 ± 2.1 |
| Embryo-level (N = 631 zygotes) |  |
| IVF fertilization, n (%) | 412 (65.3) |
| ICSI fertilization, n (%) | 219 (34.7) |
| Day 2 cell count, mean ± SD | 3.8 ± 0.7 |
| Day 3 cell count, mean ± SD | 7.4 ± 1.6 |
| Blastocyst formation (Day 5/6), n (%) | 284 (45.0) |
| Good-quality blastocyst (≥3BB), n (%) | 178 (28.2) |

**Table 2.** Comparison of key time-lapse embryo databases.

| Characteristic | This Study | Gomez et al. 2022 |
| --- | --- | --- |
| Embryos (N) | 631 | 704 |
| Cycles (N) | 218 | N/R |
| Image frames | ~500K (estimated) | 2.4M |
| Focal planes | 7 | 7 |
| Annotation type | Clinical outcomes + grades | Developmental stage labels (16 phases) |
| Clinical outcomes | Blastocyst formation (Yes/No) | None |
| Conventional grades | Day 2/3/5 Istanbul consensus | None |
| Zygote morphokinetics | 84 features | Not extracted |
| Public availability | On request | Fully open (Zenodo, CC-BY) |
| Primary use case | Prediction model development | DL model training/benchmarking |

**Table 3.** Ablation experiment results comparing model configurations.

| Model | Features | AUC | Brier Score | Sensitivity |
| --- | --- | --- | --- | --- |
| Conventional-only | 8 | 0.65 (0.60–0.70) | 0.218 | 0.52 |
| Morphokinetics-only | 16† | 0.71 (0.66–0.76) | 0.195 | 0.61 |
| Full fusion model | 23† | **0.78 (0.74–0.82)** | 0.172 | 0.68 |

†After RFE. AUC values are mean (95% CI) from nested 5-fold cross-validation.

The 45.0% blastocyst formation rate is consistent with published rates from centers employing similar culture protocols and patient demographics [7, 8].

### Feature Distribution and Quality Control Metrics

Of the 84 morphokinetic parameters extracted per zygote, 78 (92.9%) were successfully computed for all 631 embryos. The six features with partial completeness (70–95% complete) were predominantly pronuclear dynamics features whose computation required clear visualization of both pronuclei throughout the pre-syngamy period; embryos with transient PN obstruction by cytoplasmic debris accounted for these missing values. Missing morphokinetic values were imputed using the median of the feature distribution within the same fertilization-method stratum (IVF vs. ICSI).

Conventional embryo assessment features were 100% complete for Day 2 and Day 3 observations and 100% complete for blastocyst outcome labeling. The intra-class correlation coefficient between the two embryologists who performed data verification exceeded 0.91 for all ordinal grading variables.

### Ablation Experiment Results

The integrated model (GBDT-Full) achieved the highest discrimination (AUC 0.78, 95% CI 0.74–0.82), significantly outperforming both the morphokinetics-only model [GBDT-Morpho, AUC 0.71, *Δ*AUC 0.07, DeLong *Z*=3.47, *P*=0.003 (Bonferroni-corrected)] and the conventional-only model [GBDT-Conv, AUC 0.65, *Δ*AUC 0.13, DeLong *Z*=5.12, *P*<0.001]. The substantial and statistically significant AUC difference between the morphokinetics-only and conventional-only models (*Δ*AUC 0.06, *Z*=3.02, *P*=0.008) further confirms that zygote morphokinetics provide more predictive information than conventional Day 2–3 assessment alone for blastocyst formation. SHAP analysis identified cytoplasmic area change rate (mean |SHAP)=0.042, positive association), zona pellucida grayscale intensity trend (mean |SHAP)=0.038, inverted U-shaped relationship), and pronuclear fading time (mean |SHAP)=0.035, negative association—shorter fading time associated with higher blastocyst probability) as the three most influential features.

The progressive improvement from conventional-only to morphokinetics-only to the full fusion model confirms that zygote morphokinetic parameters provide complementary predictive information beyond that encoded in static morphological grades. The Brier score of 0.172 for the full model indicates reasonable calibration, although further calibration refinement (e.g., isotonic or Platt scaling) may be warranted.

### Feature Importance Analysis

SHAP value analysis of the full fusion model revealed that three morphokinetic features dominated the importance ranking: (1) cytoplasmic area slope (*d A*_cyto_ / *d t* ), reflecting the rate of cytoplasmic volume change during the first 18 hours of development; (2) zona pellucida grayscale trend, capturing progressive optical changes in the glycoprotein shell; and (3) pronuclear fading time (*t* _PNF_), a well-established morphokinetic marker of developmental competence [7, 10]. The top-ranked conventional feature was Day 3 cell count (rank 7 overall), consistent with its established prognostic value [4]. The prominence of morphokinetic features in the top ranks is consistent with the ablation experiment results and supports the biological rationale that pre-cleavage dynamics encode information about oocyte quality and developmental potential that is not fully captured by cleavage-stage morphology alone.

### Database Reuse Guide

To facilitate re-analysis by other research groups, a comprehensive data dictionary is provided as Supplementary Material documenting every variable in the database. The dictionary specifies: variable name, data type, unit of measurement, valid value range, coding scheme (for categorical variables), and the temporal window over which morphokinetic parameters were computed. Researchers requesting access to the database should include a brief analytic plan with their inquiry to facilitate review by the institutional ethics committee.

## Discussion

The primary contribution of this work is the construction of a curated, standardized database that links zygote-stage morphokinetic parameters with conventional embryo assessment outcomes, and the demonstration that gradient boosting decision trees with equal-weight initialization provide an effective framework for fusing these heterogeneous data modalities.

### Database as a Community Resource

Publicly available embryo imaging datasets suitable for machine learning research remain scarce. The Gomez et al. dataset [12, 13] represents the largest public release to date, containing 704 embryo videos with morphological annotations including blastocyst expansion grade and ICM/TE classification. However, outcome labels in that dataset are limited to morphological grades at Day 5/6, whereas our database additionally contains the binary blastocyst formation outcome and patient-level clinical variables that enable stratification analyses. Conversely, the Gomez dataset includes raw video files that our database does not release publicly due to institutional data-sharing policies. The two resources are therefore complementary: the Gomez dataset supports algorithm development on raw image data, while our database supports integrative modeling of morphokinetic parameters with clinical covariates.

With 631 embryos from 218 cycles, our database size is modest compared to multi-center registries [23, 24]. However, the depth of zygote-level morphokinetic phenotyping—84 parameters spanning four biological compartments over 18 hours of continuous observation—is, to our knowledge, substantially higher than that reported in comparable single-center datasets [14, 25]. Prospective expansion to include embryos from collaborative centers is underway.

### GBDT Fusion as a Generalizable Methodology

The equal-weight initialization strategy described here addresses a general problem in biomedical data fusion: how to combine feature modalities with different measurement scales, dimensionalities, and signal-to-noise ratios without a priori assumptions about their relative importance. In the IVF context, this approach has practical appeal because it does not require embryologists to perform additional annotations beyond standard practice, yet it systematically exploits information latent in the time-lapse record that human observers may overlook.

The ablation experiment results (full model AUC 0.78 vs. conventional-only AUC 0.65, *Δ*AUC = 0.13) quantify the incremental value of zygote morphokinetics over standard grading. The magnitude of this increment is comparable to or exceeds that reported for late-cleavage morphokinetic markers in previous studies [8, 23], suggesting that zygote-stage dynamics are an underexploited source of predictive information.

Importantly, the GBDT fusion framework is not specific to embryo assessment; it generalizes to any ART application where time-series imaging data must be integrated with structured clinical records, including oocyte maturation assessment, endometrial receptivity profiling, and embryo-endometrium synchrony modeling.

### Limitations

Several limitations should be acknowledged. First, the database is derived from a single center, which may limit generalizability to populations with different demographic profiles or laboratory protocols. Multi-center validation is required before the fusion framework can be considered broadly applicable. Second, the zygote segmentation algorithm, while validated [14, 15], is proprietary (US Patent US11210494B2) and requires a Material Transfer Agreement for academic use, which may constrain independent replication of the morphokinetic parameter extraction step. Third, the study was conducted under budgetary constraints (approximately 10,000 RMB), which limited the scope of database curation (e.g., manual verification was performed by two rather than three embryologists) and precluded the use of external computational resources for more extensive hyperparameter optimization. Fourth, the TRIPOD reporting guidelines [28] were followed for transparency, but the present paper describes only model development (Type 1b); external validation is the subject of a companion study.

### Comparison with Related Methodologies

Existing approaches to embryo assessment can be broadly categorized as: (a) static morphological grading [4]; (b) morphokinetic annotation with logistic regression or decision trees [7, 8, 9]; (c) deep learning applied to raw time-lapse images [20, 21, 22]; and (d) ensemble methods combining clinical variables with morphokinetics [24, 25]. Our framework extends category (d) by: (i) focusing specifically on the understudied zygote stage rather than cleavage-stage kinetics; (ii) employing a principled equal-weight initialization scheme that avoids modality dominance; and (iii) providing a full ablation decomposition that quantifies the marginal contribution of each data modality. The LightGBM implementation [18] additionally offers computational efficiency advantages over deep learning approaches for tabular-level data, with training times on the order of seconds rather than hours.

## Conclusions

We have constructed a standardized time-lapse imaging database containing 631 2PN zygotes with 84 zygote-stage morphokinetic parameters, 8 conventional embryo assessment features, and blastocyst formation outcomes. We have further developed a GBDT ensemble fusion framework with equal-weight initialization and residual-decreasing training that effectively integrates heterogeneous embryo data types. Ablation experiments confirm that zygote morphokinetics provide complementary predictive information beyond conventional morphological grading. The database and methodology are intended to serve as a resource and blueprint for reproducible computational embryo assessment research. The clinical validation of this framework, including direct comparison with embryologist assessments, will be reported separately.

## Data Availability

NA

## Acknowledgments

This work was supported by the Guangdong Medical Science and Technology Research Fund (A2023001). The zygote segmentation algorithm is protected under United States Patent US11210494B2.

## Author Contributions

M.P.Z. conceived the study, performed database construction and computational analyses, and drafted the manuscript. J.L., D.H., C.F.Z., Y.Z., S.P.C., and C.X.L. contributed to clinical data collection, embryo annotation, and quality control procedures. All authors reviewed and approved the final manuscript.

## Competing Interests

The authors declare that they have no competing interests.

## Data Availability

The clinical and morphokinetic database is available from the corresponding author upon reasonable request, subject to institutional ethics approval. The zygote segmentation algorithm is described in US Patent US11210494B2 and is available for academic use under a Material Transfer Agreement; inquiries should be directed to M.P.Z.

## Conflict of Interest Statement

M.-P.Z. is an inventor on US Patent US11210494B2, which protects the segmentation algorithm used in this study. The patent is assigned to Guangdong Provincial People’s Hospital. All other authors declare no conflicts of interest.

## Funding

This work was supported by the Guangdong Medical Science and Technology Research Fund (Grant No. A2023001), PI: Ming-Peng Zhao.

## AI Tool Usage Statement

Generative AI tools were used for language editing and formatting assistance during manuscript preparation. All scientific content, data analysis, and conclusions represent the original work of the authors and were reviewed and approved by all co-authors.

## Notes

### Competing Interest Statement

The authors have declared no competing interest.

### Author Declarations

Approved by the Institutional Review Board of Guangdong Provincial People's Hospital; written informed consent obtained

### Summary of Updates

This revised version has been updated as follows: Author list updated. A new corresponding author, Dr. Kubi Appiah (MD, PhD, Division of Nursing Education, School of Continuing Education, Hong Kong Baptist University), has been added as a second corresponding author (second-to-last position). The corresponding author email field now lists both authors. Abstract revised. The abstract now states that the standardized time-lapse imaging database was built upon the corresponding author's prior research groundwork, comprising 631 two-pronuclear (2PN) zygotes from 218 treatment cycles at Guangdong Provincial People's Hospital (2020-2023). Wording was tightened for clarity without changing any results. Methods clarified. The Study Population subsection now explicitly describes the prior research groundwork on zygote morphokinetic assessment, including the previously established high-precision segmentation approach (US Patent US11210494B2) and the associated 2PN zygote cohort, with the standardized database continuously refined within the present project. Imaging protocol, inclusion and exclusion criteria, and quality control procedures are described more explicitly. Figure 1 redesigned. The schematic overview of the database construction and GBDT ensemble fusion framework was rebuilt from a static image into a structured table-and-arrow layout for improved legibility and alignment. The caption was expanded to describe the five-step pipeline: database construction; zygote segmentation using the CNN algorithm (US Patent US11210494B2); extraction of 84 morphokinetic time-series features; GBDT ensemble fusion with equal-weight initialization and residual-decreasing training; and blastocyst development potential prediction (AUC 0.78). Equations numbered. The four equations in Section 2.3.2 are now numbered (1)-(4), and the in-text reference to the equal-weight initialization equation was corrected. References revised. (a) DOIs added to 13 entries. (b) Citation numbering re-sequenced so all in-text citations map correctly to the reference list; one unused reference removed. (c) Reference list presented under a proper References heading; a stray numbering artifact removed. Tables renumbered. The three data tables are now numbered consecutively (Table 1: demographic and clinical characteristics; Table 2: comparison of key time-lapse embryo databases; Table 3: ablation results), with in-text references updated. Formatting cleanup. Removed a leftover cross-reference artifact, fixed table caption styling, and made minor editorial corrections. No results, data, figures, or scientific conclusions have been altered. All authors have reviewed and approved this revised version.

